# Association between sleep quality and left ventricular structure in the Southall and Brent REvisited (SABRE) tri-ethnic study

**DOI:** 10.64898/2026.04.07.26349436

**Authors:** Eshaan Ghei, Nish Chaturvedi, Chloe Park, Alun D. Hughes, Victoria Garfield

**Affiliations:** Department of Population Science & Experimental Medicine, Institute of Cardiovascular Science, University College London; Department of Pharmacology and Therapeutics, University of Liverpool

**Keywords:** sleep, left ventricle, hypertrophy, remodelling

## Abstract

**Background:** Poor sleep quality is associated with increased cardiovascular risk, although its relationship with left ventricle (LV) structure is poorly understood and ethnic differences in the relationship between sleep and LV structure have not been studied. We investigated the association between poor sleep quality and LV structure in a tri-ethnic cohort.

**Methods:** A total of 1284 participants were analysed from the Southall and Brent Revisited (SABRE) study (age=49.9±6.2y; male 75.9%, Europeans (EU)=615, South Asians (SA)=457, African/African-Caribbean (AC)=212). A composite sleep quality score was calculated, and LV structure was measured using echocardiography. Associations between sleep quality and LV mass indexed to height^1.7^ (LVMi), relative wall thickness (RWT) and LV end-diastolic volume indexed to height^1.7^ (LVEDVi) were estimated using multivariable linear regression with adjustment for demographic and lifestyle factors across three models. Analyses were performed in the whole cohort and stratified by ethnicity.

**Results:** Compared with those who reported very good sleep quality, participants with poorer sleep quality had higher LVMi (4.8 (95% CI 1.4; 8.2)g/(m^1.7^⋅unit sleep score); *p*=0.006).

When stratifying by ethnicity, the association between sleep quality and LVMi was unconvincing in EU (1.9(-3.5, 7.3)g/(m^1.7^⋅unit sleep score); *p*=0.493), whereas poor sleep was associated with higher LVMi in AC and SA participants (9.1(1.3;16.8)g/(m^1.7^⋅unit sleep score); *p*=0.023 and 5.8(0.5;11.0)g/(m^1.7^⋅unit sleep score); *p*=0.031 respectively).

**Conclusions:** Poor sleep quality is associated with higher LVMi in older African/African-Caribbeans and South Asians, but not in Europeans. This may contribute to cardiovascular risk.

**Graphical Abstract:** 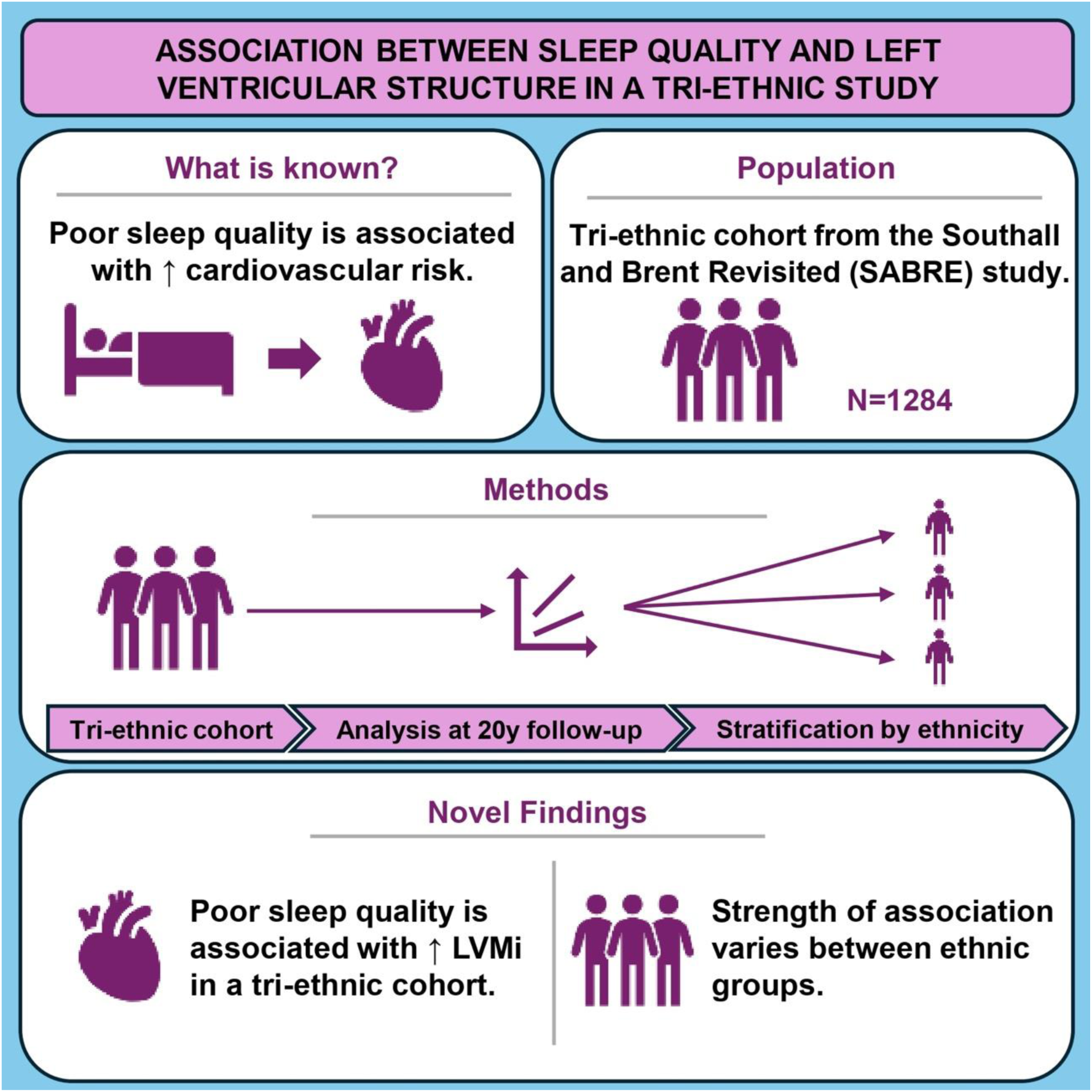

## 1. Introduction

The importance of good sleep quality for health is well documented.^1^ There is evidence that shorter and longer sleep durations are both associated with greater all-cause mortality and cardiovascular disease (CVD).^2,3^ Structural assessment of the left ventricle (LV) is important as it provides insights into LV remodelling and cardiac target organ damage; LV remodelling, especially LV hypertrophy, has been shown to be an independent predictor of CVD events.^4^ With the exception of obstructive sleep apnoea (OSA)^5^, the relationship between sleep quality and LV structure and function has not been studied extensively. Furthermore, current findings are inconsistent, and studies have included only one ethnic group,^6–10^ despite clear evidence of ethnic differences in CVD prevalence and mortality.^11,12^ Evidence also suggests variation in the association between sleep quality and CVD mortality in different ethnic groups.^13^. These findings highlight the importance of examining relationships between sleep, a potentially modifiable exposure, and cardiac structure across different ethnicities. Consequently, we investigated possible associations between sleep quality and LV structure in three ethnic groups using longitudinal data from the Southall and Brent REvisited (SABRE) study.

We hypothesized that:

1. Poor sleep quality at baseline would be associated with adverse LV structure at follow-up, independent of possible confounders.
2. Strength of associations would vary between European (EU), African/African-Caribbean (AC) and South Asian (SA) individuals.

## 2. Methods

### 2.1 Study sample

The Southall and Brent REvisited (SABRE) study is a community-based cohort of adults resident in north and west London, comprised of individuals of European, African/African-Caribbean, and South Asian ethnic backgrounds.^14,15^ The original aim of the study was to investigate possible ethnic differences in cardiometabolic risk, and by design, the study aimed to recruit more men than women.^16^ Participants of South Asian or African/African-Caribbean ethnicity were exclusively first-generation migrants and ethnicity was assigned based on participants’ parental origin.^14^ Participants gave informed consent. Ethical approval for the SABRE Visit 1 studies was granted by University College London and Ealing, Hounslow and Spelthorne, Parkside research ethics committees (EC2293). The follow-up study in 2008-11 (Visit 2) was granted ethical approval by the St Mary’s Hospital Research Ethics Committee (REC 07/H0712/109).^17^

### 2.2 Investigations

Baseline clinical investigations (visit 1) were performed in participants consecutively in Southall (1988-90) and Brent (1990-91).^14^ The combined cohort was subsequently followed up in 2008-12 (visit 2), with a median follow-up of 19 years.^17^

In addition to completing a health and lifestyle questionnaire, four indicators of sleep quality, adapted from the validated Jenkins Sleep Questionnaire, were measured at visit 1: early morning waking, difficulty falling asleep, tiredness upon waking, and snoring. This method is accepted as a valid tool for quantifying poor sleep quality without the need for specialist equipment or major inconvenience to participants.^17–19^ To reduce potential multicollinearity and comprehensively reflect the relative importance of each exposure, a composite score was calculated using principal component analysis, as previously published by Topriceanu *et al.* ^19^

At visit 2, participants attended clinic having fasted and refrained from alcohol, smoking, and caffeine for 12 or more hours before attendance. ^20^ Participants completed a questionnaire, which identified health behaviours, medical history, and medication.^20^ Smoking was categorized as never, ex- or current; alcohol consumption was categorised as none, ≤14 units per week, and >14 units per week, and years of education was used as a socioeconomic indicator due to limitations of occupational classification in multiethnic samples. Height and weight were also measured at this visit.^14^ Echocardiography was performed by 2 experienced technicians using a Philips iE33 ultrasound machine equipped with a 5.0-1.0 phased array transducer (S5-1).^20^ Calculations of LV mass, LV end diastolic volume and relative wall thickness (RWT) were guided by American Society of Echocardiography (ASE) guidelines from two-dimensional (2D) guided M-mode. ^21^ LV mass and LV end diastolic volume were indexed to height^1.7^ (LVMi and LVEDVi, respectively) to account for body size.^22^ Reproducibility of measurements was excellent as described previously.^20^

### 2.3 Statistical analysis

Frequency and percentages of categorical variables were reported alongside means and standard deviations (SD) of normally distributed continuous variables. Associations between sleep quality and outcomes were estimated using multivariable linear regression modelling, using the composite sleep score as the primary exposure variable. Associations were summarised as mean beta coefficients (95% confidence intervals). Potential confounders were selected *a priori* based on published evidence and informed by a directed acyclic graph. Care was taken not to include potential mediators such as blood pressure or diabetes in models, since they are potentially on the causal path from sleep quality to LV structure. From a conceptual standpoint, confounders were divided into two categories: demographic factors (age, sex, years of education completed and ethnicity) and lifestyle factors (smoking, alcohol consumption and BMI). The analyses were repeated using each measure of sleep quality as an independent exposure variable. Based on previous literature, it was decided *a priori* that the sample would be stratified by ethnicity and the analysis would be conducted in each sub-sample.^11–13^ The potential modifying effect of ethnicity on the association between sleep quality and LV structure was also examined by including ethnicity as an interaction term in models. For all models, a complete case analysis was performed. A flow diagram detailing participant numbers and reasons for missingness is shown in *Figure 1*. All analyses were conducted using R Studio version 4.3.0.

**Figure 1.**
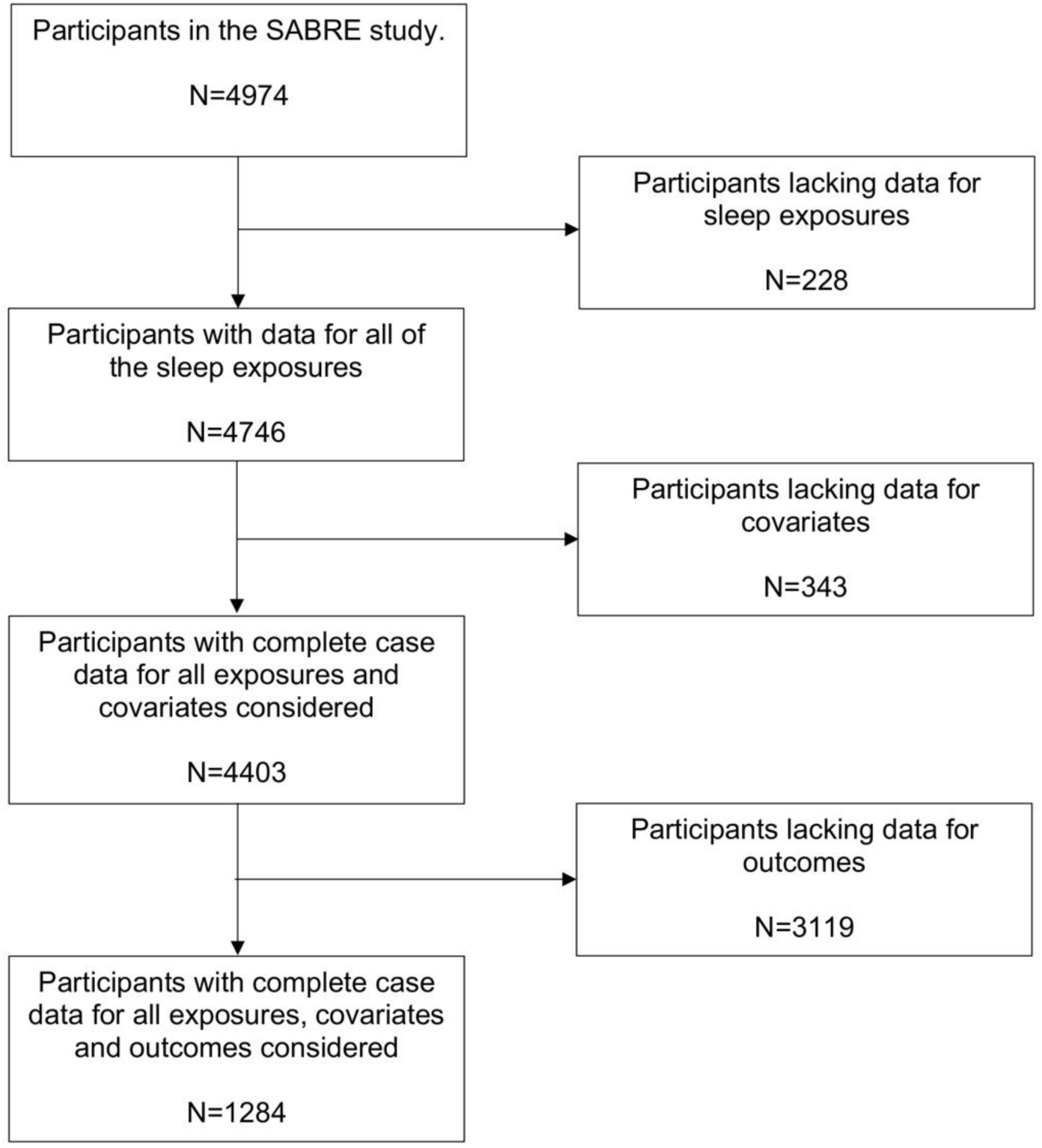
Flow diagram illustrating the derivation of the final cohort for complete case analysis.

## 3. Results

### 3.1 Sample Characteristics

Sample characteristics are summarised in *Table 1.* The mean (SD) age was 49.9 (6.2) years and participants were predominantly male. The largest proportion of participants were European (47.9%), while South Asian (35.5%) and African/African-Caribbean (16.6%) participants made up smaller proportions of the cohort. A larger proportion of South Asian participants reported early morning waking (46.8%) than European (35.4%) and African/African-Caribbean (35.4%) participants (p<0.001). The proportions of participants of each ethnicity reporting the other 3 sleep exposures were similar.

**Table 1.**
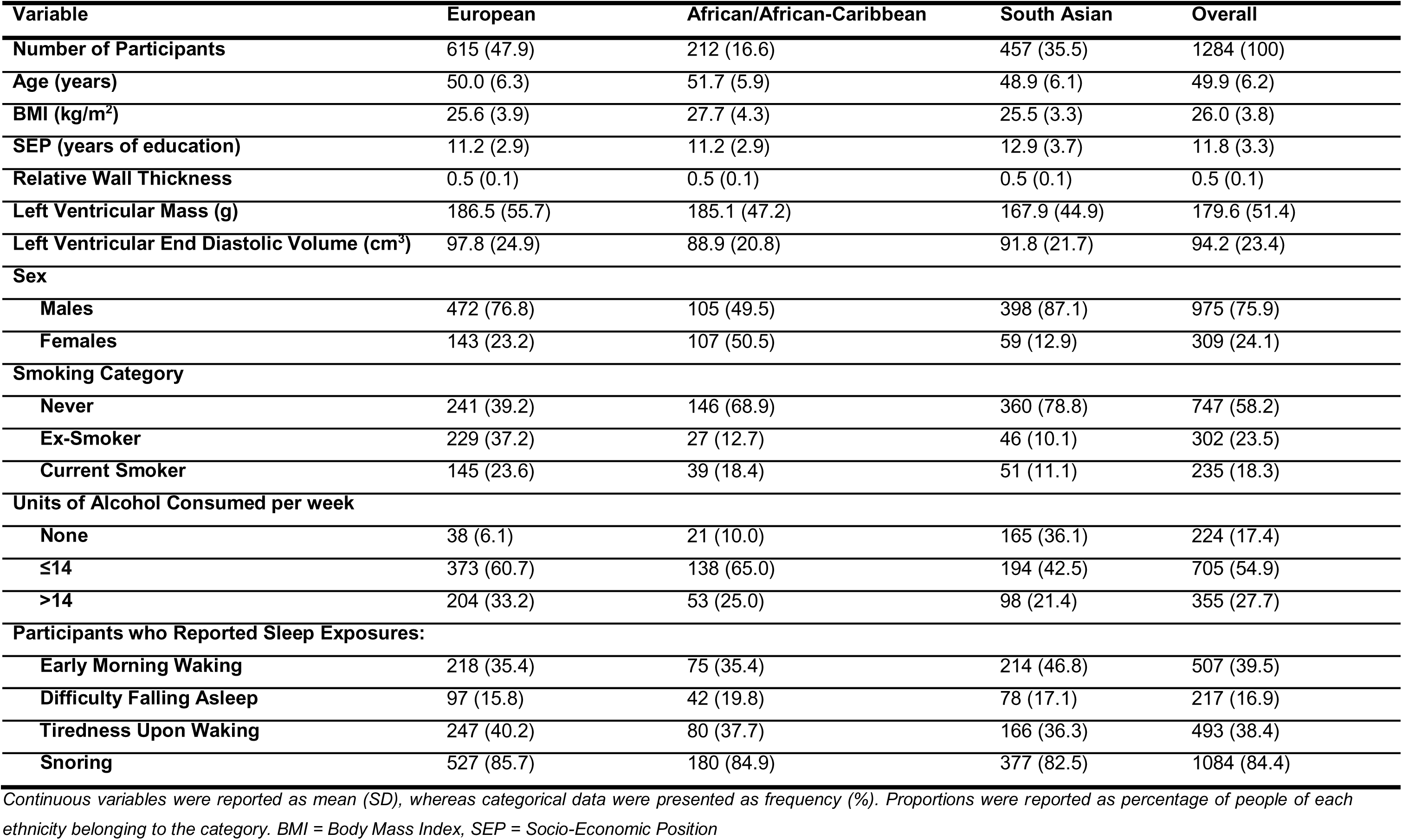
Sample characteristics of complete case cohort (N=1284), stratified by ethnicity.

| Variable | European | African/African-Caribbean | South Asian | Overall |
| --- | --- | --- | --- | --- |
| **Number of Participants** | 615 (47.9) | 212 (16.6) | 457 (35.5) | 1284 (100) |
| **Age (years)** | 50.0 (6.3) | 51.7 (5.9) | 48.9 (6.1) | 49.9 (6.2) |
| **BMI (kg/m<sup>2</sup>)** | 25.6 (3.9) | 27.7 (4.3) | 25.5 (3.3) | 26.0 (3.8) |
| **SEP (years of education)** | 11.2 (2.9) | 11.2 (2.9) | 12.9 (3.7) | 11.8 (3.3) |
| **Relative Wall Thickness** | 0.5 (0.1) | 0.5 (0.1) | 0.5 (0.1) | 0.5 (0.1) |
| **Left Ventricular Mass (g)** | 186.5 (55.7) | 185.1 (47.2) | 167.9 (44.9) | 179.6 (51.4) |
| **Left Ventricular End Diastolic Volume (cm<sup>3</sup>)** | 97.8 (24.9) | 88.9 (20.8) | 91.8 (21.7) | 94.2 (23.4) |
| **Sex** |  |  |  |  |
| **Males** | 472 (76.8) | 105 (49.5) | 398 (87.1) | 975 (75.9) |
| **Females** | 143 (23.2) | 107 (50.5) | 59 (12.9) | 309 (24.1) |
| **Smoking Category** |  |  |  |  |
| **Never** | 241 (39.2) | 146 (68.9) | 360 (78.8) | 747 (58.2) |
| **Ex-Smoker** | 229 (37.2) | 27 (12.7) | 46 (10.1) | 302 (23.5) |
| **Current Smoker** | 145 (23.6) | 39 (18.4) | 51 (11.1) | 235 (18.3) |
| **Units of Alcohol Consumed per week** |  |  |  |  |
| **None** | 38 (6.1) | 21 (10.0) | 165 (36.1) | 224 (17.4) |
| **≤14** | 373 (60.7) | 138 (65.0) | 194 (42.5) | 705 (54.9) |
| **>14** | 204 (33.2) | 53 (25.0) | 98 (21.4) | 355 (27.7) |
| **Participants who Reported Sleep Exposures:** |  |  |  |  |
| **Early Morning Waking** | 218 (35.4) | 75 (35.4) | 214 (46.8) | 507 (39.5) |
| **Difficulty Falling Asleep** | 97 (15.8) | 42 (19.8) | 78 (17.1) | 217 (16.9) |
| **Tiredness Upon Waking** | 247 (40.2) | 80 (37.7) | 166 (36.3) | 493 (38.4) |
| **Snoring** | 527 (85.7) | 180 (84.9) | 377 (82.5) | 1084 (84.4) |

Continuous variables were reported as mean (SD), whereas categorical data were presented as frequency (%). Proportions were reported as percentage of people of each ethnicity belonging to the category. BMI = Body Mass Index, SEP = Socio-Economic Position

### 3.2 Associations between composite sleep score and LV structure

A unit increase in the composite sleep score (i.e. worse sleep quality) was associated with higher LVMi (β = 5.2 (95% CI 1.3, 9.0) g/(m^1.7^⋅unit sleep score)) in the unadjusted model (Model 1, *Fig. 2A*). The association increased slightly after adjustment for demographic factors (Model 2) and was slightly attenuated to 4.8 (1.4, 8.2)g/(m^1.7^⋅unit sleep score) (p=0.006) after full adjustment (Model 3). There was no convincing association between the composite sleep score and LVEDVi, in any models although the estimates had wide 95% confidence intervals (*Fig. 2B*). There was no evidence to support an association between the composite sleep score and RWT (*Fig. 2C)*.

**Figure 2.**
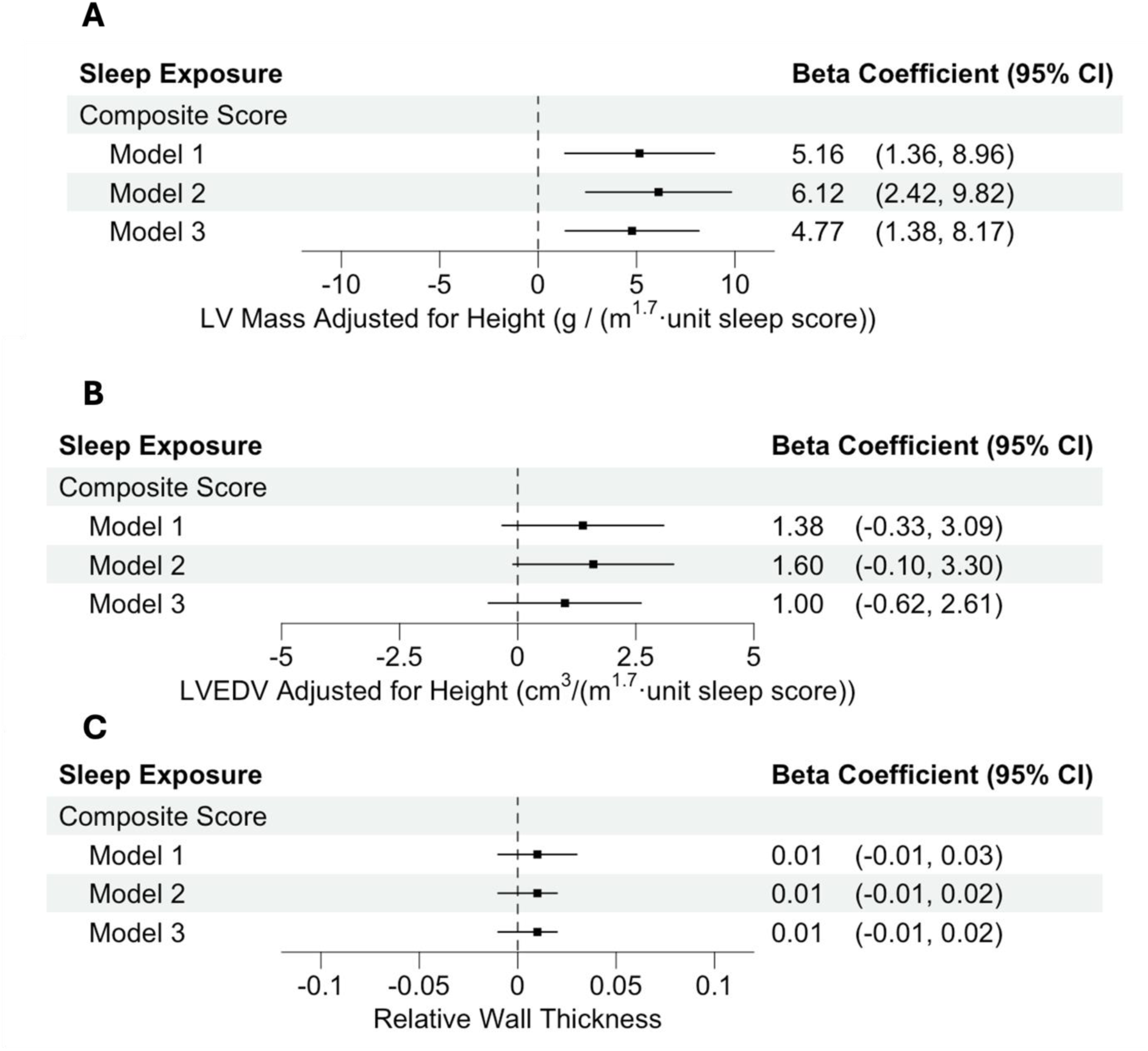
Forest Plots showing associations between composite sleep score and A) LV mass adjusted for height B) LVEDV adjusted for height and C) Relative Wall Thickness. Model 1 = Unadjusted univariate analysis. Model 2 = Adjusted for demographic factors (age, sex, ethnicity, years of education). Model 3 = Fully adjusted model including adjustments for lifestyle factors (BMI, smoking, alcohol consumption). N=1284. All results were obtained by linear regression analysis, using R studio version 4.3.0. 95% CI = 95% Confidence Interval, LV = Left Ventricle, EDV = End Diastolic Volume.

### 3.3 Associations between individual sleep quality exposures and LVMi, LVEDVi and RWT

Snoring was associated with a higher LVMi in Model 1 and Model 2 (*Fig. 3A*). However, when further adjusted for lifestyle factors in Model 3, the association diminished to 1.4(-1.4, 4.2) g/(m^1.7^⋅unit snoring score) (*p*=0.34), i.e. compatible with no association. In contrast, early morning waking was associated with higher LVMi in all 3 models; the size of this association was 3.6 (1.5, 5.7) g/(m^1.7^⋅unit sleep score) (p<0.001) for the fully adjusted model. There were no convincing associations between difficulty in falling asleep or tiredness upon waking and LVMi (*Fig 3A)*.

**Figure 3.**
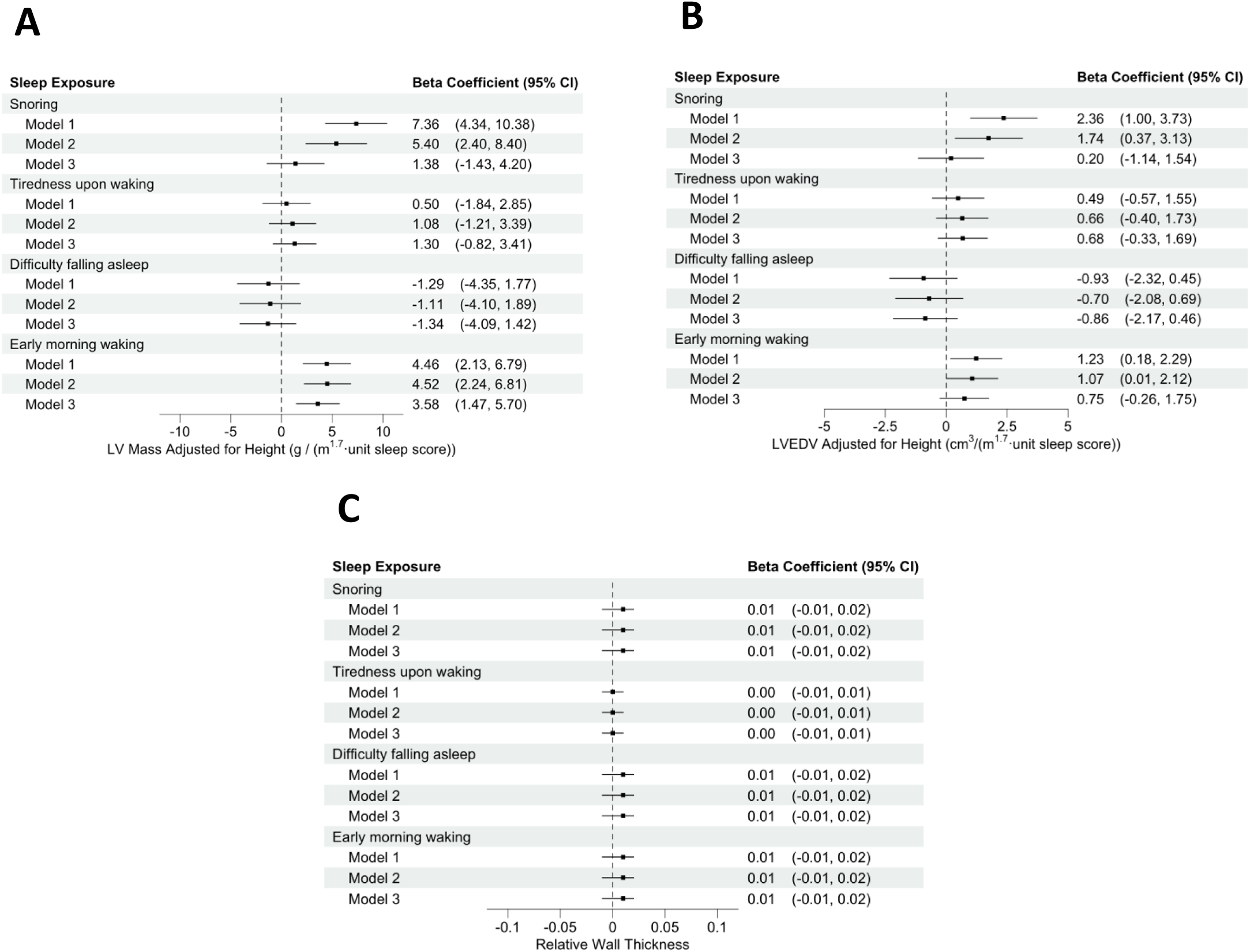
Forest Plots showing associations between the simultaneously inputted sleep exposures and A) LV mass adjusted for height B) LVEDV adjusted for height and C) Relative wall thickness. Model 1 = Unadjusted univariate analysis. Model 2 = Adjusted for demographic factors (age, sex, ethnicity, years of education). Model 3 = Fully adjusted model including adjustments for lifestyle factors (BMI, smoking, alcohol consumption). N=1284. All results were obtained by linear regression analysis, using R studio version 4.3.0. 95% CI = 95% Confidence Interval, LV = Left Ventricle, EDV = End Diastolic Volume.

Snoring was associated with a higher LVEDVi in Model 1 and Model 2 but not convincingly in the fully adjusted Model 3 (*Fig. 3B).* Similarly, early morning waking was associated with higher LVEDVi in Model 1 and Model 2, but this association was attenuated after full adjustment (Model 3: 0.8 (-0.3, 1.8)cm^3^/(m^1.7^⋅unit sleep score), *p*=0.15). There were no convincing associations between difficulty in falling asleep or tiredness upon waking and LVEDVi (*Fig 3B)*.

No association was seen between any of the sleep quality exposures and RWT (*Fig. 3C)*.

### 3.4 Associations between composite sleep score and LVMi, LVEDVi and RWT, stratified by ethnicity

There were no convincing associations in any of the three models between sleep score and LVMi in the Europeans (*Fig. 4A)*. However, an association was observed for all three models in participants of African/African-Caribbean ethnicity; in the fully adjusted model, this corresponded to an increase of 9.1 (1.3, 16.8) g/(m^1.7^⋅unit sleep score) (p=0.023) (*Fig 4A)*. A similar, albeit slightly weaker, association between sleep score and LVMi was also observed in South Asians (5.8 (0.5, 11.0) g/(m^1.7^⋅unit sleep score); p=0.031). There was no statistically significant interaction between ethnicity and the association between sleep score and LVMi (p=0.211 for African/African-Caribbeans, p=0.472 for South Asians).

**Figure 4.**
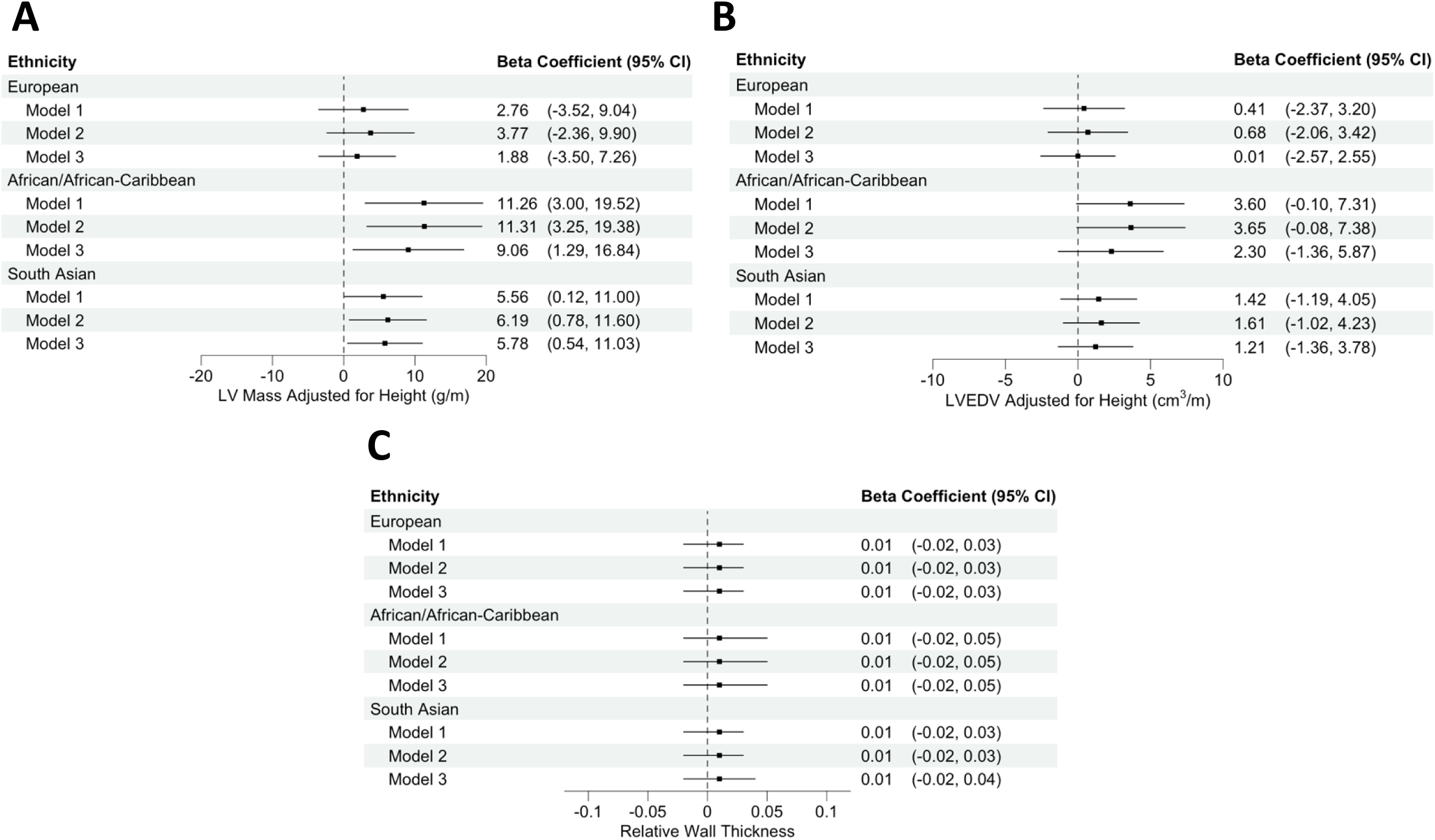
Forest Plots showing associations in samples stratified by ethnicity between the composite sleep score and A) LV mass adjusted for height B) LVEDV adjusted for height and C) Relative wall thickness. Model 1 = Unadjusted univariate analysis. Model 2 = Adjusted for demographic factors (age, sex, ethnicity, years of education). Model 3 = Fully adjusted model including adjustments for lifestyle factors (BMI, smoking, alcohol consumption). N=1284. All results were obtained by linear regression analysis, using R studio version 4.3.0. 95% CI = 95% Confidence Interval, LV = Left Ventricle, EDV = End Diastolic Volume.

Associations between sleep score and LVEDVi were close to zero in Europeans (*Fig. 4B)* and associations were not convincingly different from zero for the two other ethnic groups.

No associations between sleep score and RWT were seen after stratification by ethnicity. (*Fig. 4C)*.

## 4. Discussion

This study used a tri-ethnic UK population-based sample to investigate the association between sleep quality and LV structure, as measured by LVMi, LVEDVi and RWT. Overall, poorer sleep quality was associated with greater LVMi at 20-year follow-up, with or without adjustment for potential confounders. The strongest predictor for a higher LVMi was early morning waking. There was no convincing association between sleep quality and LVEDVi or RWT after full adjustment for confounders. In analyses stratified by ethnicity, associations between poor sleep quality and higher LVMi were stronger in participants of African/African-Caribbean and South Asian ethnicity.

As far as we are aware, there have been no previous studies looking at the association between sleep quality and LV structure in a multi-ethnic cohort. Previous studies in Norway^7,8^ and Korea^9^ have provided conflicting evidence on the relationship between sleep quality or duration and LV structure and function. Our data suggest that ethnic differences in these relationships could contribute to these inconsistent findings. Stronger associations between poor sleep quality and higher LVMi were observed in participants of African/African-Caribbean and South Asian ethnicity. South Asians have a higher risk of most forms of CVD compared with Europeans in the UK,^11,12^ while African/African-Caribbean people in UK are at lower risk of coronary artery disease, but at higher risk of stroke.^11,12^ Both ethnic groups experience a higher burden of heart failure.^23^ Our findings provide a plausible mechanism to link poor sleep quality to these adverse outcomes, but this remains to be established.

Our observation that poorer sleep quality was associated with higher LVMi accompanied by a normal RWT is characteristic of eccentric hypertrophy.^24,25^ Evidence that chamber volume increased was equivocal, but this may reflect a lack of precision in estimating LVEDV using 2D echocardiography.^26^ The possible mechanisms by which poor sleep may induce pathological cardiac remodelling may include activation of inflammatory pathways^27^; activation of the sympathetic nervous system and renin-angiotensin-aldosterone system^28–30^, or alterations in glucose and lipid metabolism.^31^ It is noteworthy that both diabetes^32^ and hypertension^33^ are more common in older people of African/African-Caribbean and South Asian ethnicity in the UK. The potential role for diabetes and hypertension, especially nocturnal blood pressure, as mediators of the association between poor sleep quality and LV hypertrophy should be examined in future studies.

When sleep exposures were examined separately, associations were found between early morning waking and snoring with both LVMi and LVEDVi. Early morning waking is typically indicative of sleep maintenance insomnia and, by extension, reduced quality of sleep.^34^ Patients reporting early morning waking are also more likely to report low sleep duration.^34^ Sleep maintenance insomnia becomes more common with advancing age due to less time spent in the deeper stages of sleep,^35^ but these associations were independent of age and hardly attenuated by adjustment for other confounders. Snoring is regarded as a reliable proxy marker for OSA.^36^ Associations between snoring and LVMi were largely attenuated after adjustment, implying that the relationship between snoring and LV structure is confounded, particularly by BMI.

This study possesses a number of strengths and limitations. Firstly, the use of the SABRE cohort allowed for inclusion of a wide range of potential confounders; however, the possibility of residual confounding cannot be excluded. The extended follow-up period of 20 years allowed for cardiac remodelling to take place.^14^ Furthermore, the use of echocardiography permitted accurate and cost-effective estimations of LV structure, despite lacking the precision of cardiac MRI.^37,38^ 2D echocardiography was used in this study due to relative unavailability of, and hence incomplete data for, 3D echocardiography in visit 2 of the SABRE study. A limitation of this study was the use of self-reported sleep quality. This is less accurate than objective measures, such as actigraphy and polysomnography, but at the time of baseline measurements, such measures were impractical at scale. As expected with a 20-year follow-up period, there was substantial loss to follow up; out of 4974 people in the cohort at baseline, only 1284 had echocardiography data. As described previously,^39^ people attending for echocardiography tended to be healthier than those that did not and this will have introduced selection bias; however, this might be anticipated to attenuate observed relationships. Another limitation is that when stratifying by ethnicity, sample sizes were reduced, and statistical power was diminished. This was particularly true when considering the African/African-Caribbean cohort. This will have limited the precision of our estimates of the association in the stratified analyses.

### Perspectives

In conclusion, self-reported poor sleep quality was found to be associated with a greater LVMi in a UK tri-ethnic cohort. Associations between composite sleep score and LVMi were stronger in South Asian and African/African-Caribbean people. Further research is required to determine the implications of these structural changes, but these findings highlight the potential for including methods of improving sleep quality in the management of patients at risk of CVD.

### Novelty and Relevance

#### What is new?

Poor sleep quality is associated with greater left ventricular (LV) mass at follow up in a tri-ethnic cohort consisting of people of European, South Asian and African/African-Caribbean ethnicity in the UK.

#### What is relevant?

Poor sleep is associated with cardiovascular disease (CVD). Left ventricular hypertrophy is an indicator of increased CVD risk. People of South Asian and African/African-Caribbean ethnicity in the UK are at increased risk of CVD and stroke respectively.

#### Clinical and pathophysiological perspectives

Pathophysiological remodelling of the left ventricle may contribute to the known association between poor sleep and increased incidence of cardiovascular events. Understanding this association and its ethnic differences might provide a potential target for intervention.

## Data Availability

SABRE data is available to approved researchers upon reasonable request. Details can be found at: https://www.sabrestudy.org/home-2/datasharing/.

## Non-standard Abbreviations and Acronyms

2D: Two-dimensional
AC: African/African-Caribbean
CVD: Cardiovascular disease
EU: European
LV: Left ventricle
LVEDVi: Left ventricular end diastolic volume indexed to height^1.7^
LVMi: Left ventricular mass indexed to height^1.7^
OSA: Obstructive sleep apnoea
RWT: Relative wall thickness
SA: South Asian
SABRE: Southall and Brent REvisited
SD: Standard deviation

## Acknowledgements

We are extremely grateful to all the people who took part in the study, and past and present members of the SABRE team who helped collect the data.

## References

1. Nelson, K. L., Davis, J. E. & Corbett, C. F. Sleep quality: An evolutionary concept analysis. Nurs. Forum (Auckl). 57, 144–151 (2022).

2. Yin, J. et al. Relationship of Sleep Duration With All-Cause Mortality and Cardiovascular Events: A Systematic Review and Dose-Response Meta-Analysis of Prospective Cohort Studies. J. Am. Heart Assoc. 6, (2017).

3. Kwok, C. S. et al. Self-Reported Sleep Duration and Quality and Cardiovascular Disease and Mortality: A Dose-Response Meta-Analysis. J. Am. Heart Assoc. 7, (2018).

4. Levy, D., Garrison, R. J., Savage, D. D., Kannel, W. B. & Castelli, W. P. Prognostic implications of echocardiographically determined left ventricular mass in the Framingham Heart Study. N. Engl. J. Med. 322, 1561–6 (1990).

5. Liu, L. et al. Obstructive Sleep Apnea and Hypertensive Heart Disease: From Pathophysiology to Therapeutics. Rev. Cardiovasc. Med. 24, 342 (2023).

6. Ingelsson, E., Lind, L., Ärnlöv, J. & Sundström, J. Sleep disturbances independently predict heart failure in overweight middle-aged men. Eur. J. Heart Fail. 9, 184–190 (2007).

7. Laugsand, L. E., Strand, L. B., Platou, C., Vatten, L. J. & Janszky, I. Insomnia and the risk of incident heart failure: a population study. Eur. Heart J. 35, 1382–1393 (2014).

8. Strand, L. B., Laugsand, L. E., Dalen, H., Vatten, L. & Janszky, I. Insomnia and left ventricular function – an echocardiography study. Scandinavian Cardiovascular Journal 50, 187–192 (2016).

9. Lee, J.-H. et al. Sleep Duration and Quality as Related to Left Ventricular Structure and Function. Psychosom. Med. 80, 78–86 (2018).

10. Krokstad, S. et al. Cohort Profile: The HUNT Study, Norway. Int. J. Epidemiol. 42, 968–977 (2013).

11. George, J. et al. Ethnicity and the first diagnosis of a wide range of cardiovascular diseases: Associations in a linked electronic health record cohort of 1 million patients. PLoS One 12, e0178945 (2017).

12. Razieh, C. et al. Differences in the risk of cardiovascular disease across ethnic groups: UK Biobank observational study. *Nutrition*, Metabolism and Cardiovascular Diseases 32, 2594–2602 (2022).

13. Garfield, V., Joshi, R., Garcia-Hernandez, J., Tillin, T. & Chaturvedi, N. The relationship between sleep quality and all-cause, CVD and cancer mortality: the Southall and Brent REvisited study (SABRE). Sleep Med. 60, 230–235 (2019).

14. Tillin, T., Forouhi, N. G., McKeigue, P. M. & Chaturvedi, N. Southall And Brent REvisited: Cohort profile of SABRE, a UK population-based comparison of cardiovascular disease and diabetes in people of European, Indian Asian and African Caribbean origins. Int. J. Epidemiol. 41, 33–42 (2012).

15. McKeigue, P. M., Shah, B. & Marmot, M. G. Relation of central obesity and insulin resistance with high diabetes prevalence and cardiovascular risk in South Asians. The Lancet 337, 382–386 (1991).

16. Jones, S. et al. Cohort Profile Update: Southall and Brent Revisited (SABRE) study: a UK population-based comparison of cardiovascular disease and diabetes in people of European, South Asian and African Caribbean heritage. Int. J. Epidemiol. 49, 1441–1442e (2020).

17. Ong, Z. L., Chaturvedi, N., Tillin, T., Dale, C. & Garfield, V. Association between sleep quality and type 2 diabetes at 20-year follow-up in the Southall and Brent REvisited (SABRE) cohort: a triethnic analysis. J. Epidemiol. Community Health (1978). 75, 1117–1122 (2021).

18. Jenkins, C. D., Stanton, B.-A., Niemcryk, S. J. & Rose, R. M. A scale for the estimation of sleep problems in clinical research. J. Clin. Epidemiol. 41, 313–321 (1988).

19. Topriceanu, C., Tillin, T., Chaturvedi, N., Joshi, R. & Garfield, V. The association between plasma metabolites and sleep quality in the Southall and Brent Revisited (SABRE) Study: A cross-sectional analysis. J. Sleep Res. 30, (2021).

20. Park, C. M. et al. Left-Ventricular Structure in the Southall And Brent REvisited (SABRE) Study. Hypertension 61, 1014–1020 (2013).

21. Lang, R. M. et al. Recommendations for Chamber Quantification: A Report from the American Society of Echocardiography’s Guidelines and Standards Committee and the Chamber Quantification Writing Group, Developed in Conjunction with the European Association of Echocardiography, a Branch of the European Society of Cardiology. Journal of the American Society of Echocardiography 18, 1440–1463 (2005).

22. Dewey, F. E., Rosenthal, D., Murphy, D. J., Froelicher, V. F. & Ashley, E. A. Does Size Matter? Circulation 117, 2279–2287 (2008).

23. Sosin, M. D., Bhatia, G. S., Zarifis, J., Davis, R. C. & Lip, G. Y. H. An 8-year follow-up study of acute admissions with heart failure in a multiethnic population. Eur. J. Heart Fail. 6, 669–672 (2004).

24. Sayin, B. Y. & Oto, A. Left Ventricular Hypertrophy: Etiology-Based Therapeutic Options. Cardiol. Ther. 11, 203–230 (2022).

25. Grossman, W. & Paulus, W. J. Myocardial stress and hypertrophy: a complex interface between biophysics and cardiac remodeling. Journal of Clinical Investigation 123, 3701–3703 (2013).

26. Houck, R. C., Cooke, J. E. & Gill, E. A. Live 3D echocardiography: a replacement for traditional 2D echocardiography? AJR Am. J. Roentgenol. 187, 1092–106 (2006).

27. Spagnoli, L. G., Bonanno, E., Sangiorgi, G. & Mauriello, A. Role of Inflammation in Atherosclerosis. Journal of Nuclear Medicine 48, 1800–1815 (2007).

28. Somers, V. K., Dyken, M. E., Clary, M. P. & Abboud, F. M. Sympathetic neural mechanisms in obstructive sleep apnea. Journal of Clinical Investigation 96, 1897–1904 (1995).

29. Mancia, G., Grassi, G., Giannattasio, C. & Seravalle, G. Sympathetic Activation in the Pathogenesis of Hypertension and Progression of Organ Damage. Hypertension 34, 724–728 (1999).

30. Loh, H. H. et al. Influence and implications of the renin–angiotensin–aldosterone system in obstructive sleep apnea: An updated systematic review and meta-analysis. J. Sleep Res. 32, (2023).

31. Hong, S., Lee, D.-B., Yoon, D.-W., Yoo, S.-L. & Kim, J. The Effect of Sleep Disruption on Cardiometabolic Health.. 15, 60 (2025).

32. Tillin, T. et al. Insulin resistance and truncal obesity as important determinants of the greater incidence of diabetes in Indian Asians and African Caribbeans compared with Europeans: the Southall And Brent REvisited (SABRE) cohort. Diabetes Care 36, 383–93 (2013).

33. Primatesta, P., Bost, L. & Poulter, N. Blood pressure levels and hypertension status among ethnic groups in England. J. Hum. Hypertens. 14, 143–148 (2000).

34. Pavlova, M. Circadian Rhythm Sleep-Wake Disorders. CONTINUUM: Lifelong Learning in Neurology 23, 1051–1063 (2017).

35. Fiorentino, L. & Martin, J. L. Awake at 4 a.m.: treatment of insomnia with early morning awakenings among older adults. J. Clin. Psychol. 66, 1161–1174 (2010).

36. Zhou, Y. et al. Association of snoring and body composition in (peri-post) menopausal women. BMC Womens Health 20, 175 (2020).

37. Tsao, C. W. et al. Left Ventricular Structure and Risk of Cardiovascular Events: A Framingham Heart Study Cardiac Magnetic Resonance Study. J. Am. Heart Assoc. 4, (2015).

38. Margossian, R. et al. Comparison of Echocardiographic and Cardiac Magnetic Resonance Imaging Measurements of Functional Single Ventricular Volumes, Mass, and Ejection Fraction (from the Pediatric Heart Network Fontan Cross-Sectional Study)††A list of participating institutions and investigators appears in the Appendix. Am. J. Cardiol. 104, 419–428 (2009).

39. Al Saikhan, L. et al. Imaging Protocol, Feasibility, and Reproducibility of Cardiovascular Phenotyping in a Large Tri-Ethnic Population-Based Study of Older People: The Southall and Brent Revisited (SABRE) Study. Front. Cardiovasc. Med. 7, (2020).

